# Seasonal betacoronavirus antibodies expansion post BNT161b2 vaccination associates with reduced SARS-CoV-2 VoCs neutralization

**DOI:** 10.1101/2021.08.15.21262000

**Authors:** Stefania Dispinseri, Ilaria Marzinotto, Cristina Brigatti, Maria Franca Pirillo, Monica Tolazzi, Elena Bazzigaluppi, Andrea Canitano, Martina Borghi, Alessandra Gallinaro, Roberta Caccia, Riccardo Vercesi, Paul F McKay, Fabio Ciceri, Lorenzo Piemonti, Donatella Negri, Paola Cinque, Andrea Cara, Vito Lampasona, Gabriella Scarlatti

**Author notes:** Correspondence to Vito Lampasona. S. Dispinseri and I. Marzinotto contributed equally to this paper.

## Abstract

SARS-CoV-2 vaccination is known to induce antibodies that recognize also variants of concerns (VoCs) of the virus. However, epidemiological and laboratory evidences indicate that these antibodies have a reduce neutralization ability against VoCs. We studied binding and neutralizing antibodies against the Spike RBD and S2 domains of the Wuhan-Hu-1 virus and its alpha and beta VoCs and of seasonal betacoronaviruses (HKU1 and OC43) in a cohort of 31 health care workers vaccinated with BNT162b2-Comirnaty and prospectively followed post-vaccination. The study of sequential samples collected up to 64 days post-vaccination showed that serological assays measuring IgG against Wuhan-Hu-1 antigens were a poor proxy for VoCs neutralization. In addition, in subjects who had asymptomatic or mild COVID-19 prior to vaccination the loss of nAbs following disease can be rapid and protection from re-infection post-vaccination is often no better than in naïve subjects. Interestingly, in health care workers naïve for SARS-CoV-2 infection, vaccination induced a rapid and transient reactivation of pre-existing seasonal coronaviruses IgG responses that was associated with a subsequent reduced ability to neutralize some VoCs.

## Introduction

Large scale SARS-CoV-2 vaccination campaigns are ongoing though with large disparities between countries (Abecassis, 2021). While antibodies to SARS-CoV-2 are not the only determinant of immunity, they are important predictors of immunological protection from SARS-CoV-2 infection or disease (Grifoni et al., 2020). However, the continuous worldwide emergence of SARS-CoV-2 variants of concern (VoC) is challenging the effectiveness of licensed vaccines with several open questions remaining about the effectiveness and persistence of vaccine induced antibodies (Harvey et al., 2021; Cohen, 2021). Public health policies need to adapt on rolling basis and several governments are already introducing additional rounds of vaccination in subjects at risk of severe disease (High Demand for the Third COVID Vaccine Dose: More than a Quarter of a Million People Have Already Been Vaccinated). Moreover, the contribution to community immunity of subjects who experienced an asymptomatic or mild COVID-19 is unclear.

To address some of these issues, in this study we analyzed prospectively the antibody response to SARS-CoV-2 and seasonal betacoronaviruses induced by BNT162b2-Comirnaty vaccination in a cohort of health care workers (HCW) with or without a prior COVID-19. Overall, our results suggest that: 1) vaccine induced Wuhan-Hu-1 RBD IgGs should be used with caution as proxy for VoCs neutralization, 2) in subjects who had asymptomatic or mild COVID-19, the loss of neutralizing antibodies (nAbs) following disease can be rapid and post-vaccination protection from re-infection mediated by antibodies is often no better than in naïve subjects, and 3) the reactivation of pre-existing seasonal coronaviruses antibody responses can be associated with a reduced ability of the BNT162b2-Comirnaty vaccine to induce nAbs against some VoCs.

## Results and Discussion

We used validated lentiviral vector-based SARS-CoV-2 neutralization and Luciferase Immuno Precipitation System assays (Dispinseri et al., 2021; Secchi et al., 2020) to profile the antibody response to spike antigens of SARS-CoV-2 (Wuhan-hu-1, alpha and beta VoCs) and of seasonal betacoronaviruses (HKU1 and OC43) post BNT162b2-Comirnaty vaccination (Supplemental Figure 1). We tested sera from 31 vaccinated HCW of the IRCCS Ospedale San Raffaele, Milan, Italy, with (n=18) or without (n=13) prior confirmed COVID-19 (Supplemental Table 1). All but two subjects received two vaccine doses 21 days apart, between January 7^th^ and March 5^th^, 2021. We collected serum samples prospectively at first vaccination (baseline), and thereafter at 10, 21, 31, 42, and 64 days.

### Binding and neutralizing antibodies against Wuhan-Hu-1 antigens in BNT162b2 vaccinees stratified by SARS-CoV-2 antibody status at baseline

At baseline before vaccination, SARS-CoV-2 Wuhan-Hu-1 nAbs and IgGs binding to spike RBD were absent in 6 out of 18 HCW with a prior history of COVID-19 and in all those without. All vaccinees responded to BNT162b2-Comirnaty immunization by producing antibodies able to both neutralize SARS-CoV-2 Wuhan-Hu-1 and bind its RBD and S2 domains (Supplemental Figure 2, A-C). Antibody binding to the Wuhan-Hu-1 spike RBD and S2 domains and neutralization titer increased above assay thresholds starting from day 10 post-vaccination. We observed a peak of antibody responses already at day 10 in HCW with prior COVID-19 and detectable SARS-CoV-2 antibodies at baseline. In HCW naïve for COVID-19 or without SARS-CoV-2 antibodies at baseline, the antibody response peaked instead at day 31, i.e. 10 days after the booster jab. Peak values were higher in HCW who presented with SARS-CoV-2 antibodies at baseline for both nAbs (ID50 geometric mean titer to Wuhan-Hu-1 at day 31: in baseline antibody positive HCW: = 13613; in baseline antibody negative HCW = 1607) and IgG to RBD and S2 spike domains (Supplemental Table 2).

### Post-vaccination, the antibody response shows significant differences in nAb titer but not in IgG binding to SARS-CoV-2 variants of concern

nAbs and RBD IgGs against SARS-CoV-2 alpha and beta VoCs had post-vaccination kinetics similar to that against Wuhan-Hu-1, with delayed and lower peak titers in COVID-19 vaccinees without detectable antibodies at baseline (Figure 1, A and B). nAb peak titers against VoCs were reduced compared with nAbs to Wuhan-Hu-1. The reduction of peak VoC beta nAb titer was statistically significant both in HCW naïve for COVID-19 or HCW who presented with SARS-CoV-2 antibodies at baseline (Figure 1A) (ID50 geometric mean titer at day 31: in baseline antibody positive HCW: Wuhan-Hu-1 = 13613, alpha = 11862, beta = 3211; in baseline antibody negative: 1607, 801, 238, respectively).

**Figure 1.**
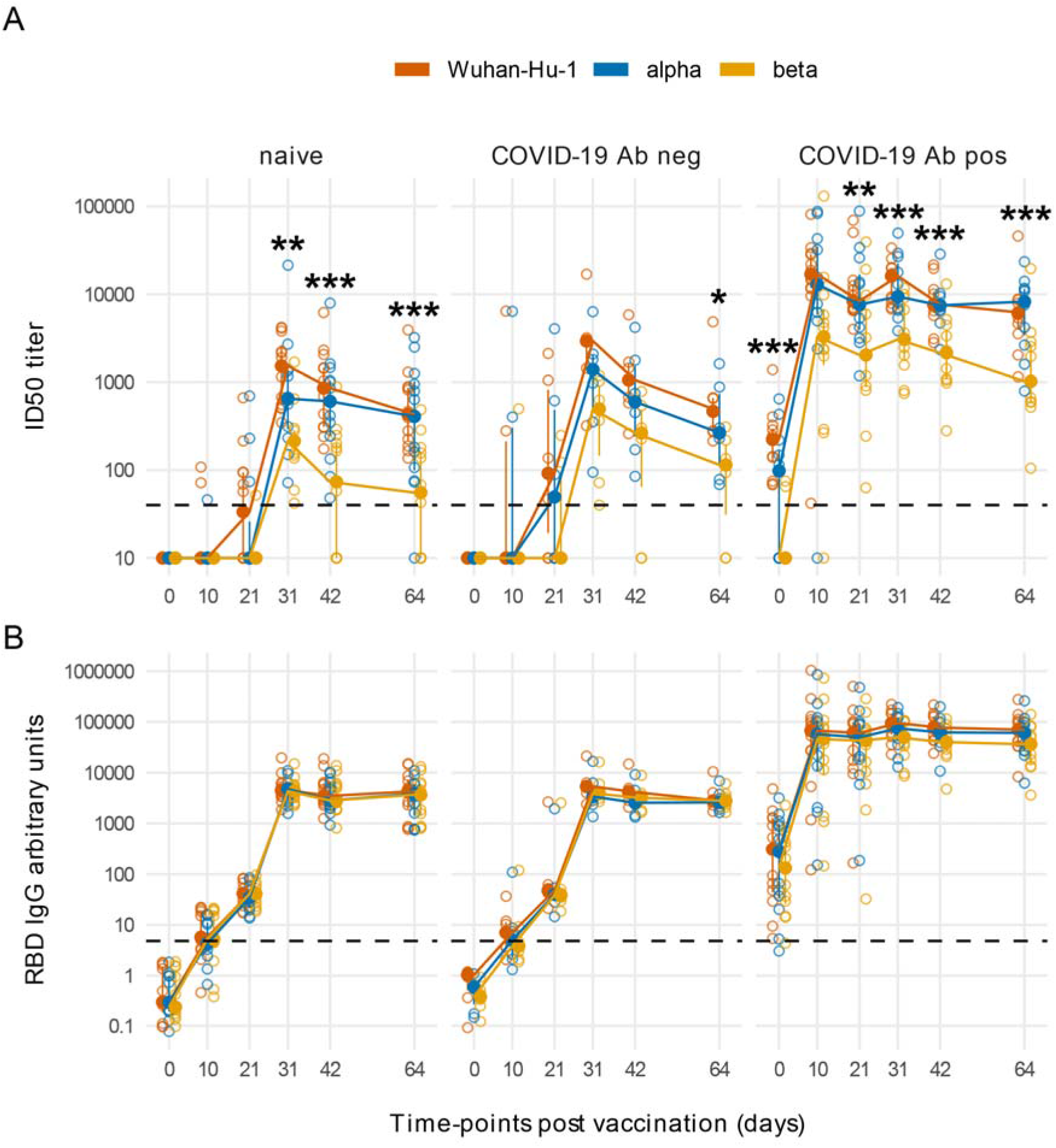
Antibody responses to Spike antigens of SARS-CoV-2 variants post BNT162b2 vaccination show significant differences in nAbs titer but not in IgG binding to the RBD of VoCs. Line plots of ID50 **(A)** and RBD IgG AU **(B)** against SARS-CoV-2 Wuhan-Hu-1 and VoCs in sequential samples after vaccination. Vaccinees are presented in left to right panels stratified as: 1) subjects naïve for SARS-CoV-2 infection (n=13), 2) with prior confirmed COVID-19 presenting at vaccination either without Wuhan-Hu-1 nAbs and RBD IgGs (n=6), and 3) with prior COVID-19 and SARS-CoV-2 antibodies at baseline (n=12). Filled circles and bars represent the median ± inter quartile range at each time-point, empty circles show each individual measurement. The horizontal dashed lines stand for the threshold for positivity. The asterisks indicate statistically significant differences in mean titers of nAbs or RBD IgGs levels across VoCs at the corresponding time-points (two-way repeated measures ANOVA, * p < 0.05, ** p< 0.01, *** p<0.001).

nAbs declined already at day 42 after vaccination (day 64/day 31 nAb geometric mean titer ratio: 0.32, 0.38 and 0.22 for Wuhan-Hu-1, alpha and beta, respectively) with nAbs against VoC beta dropping below the threshold for positivity in 7 HCW, all of whom were antibody negative at baseline (5 naïve for SARS-CoV-2 infection and 2 with prior COVID-19). Conversely, IgG binding to Wuhan-Hu-1 and VoC RBD domains was comparable and showed only a modest drop by day 64 (Figure 1B).

### Post-vaccination, antibody levels to spike antigens of SARS-CoV-2 and its variants are correlated to previous disease severity

HCW with prior COVID-19 were stratified according to disease severity (https://www.who.int/publications-detail-redirect/WHO-2019-nCoV-clinical-2021-1): disease was asymptomatic in 5, mild in 11 and moderate in 2 (Supplemental Table 1). After BNT162b2 vaccination, we observed that nAb peak values increased according to disease severity in HCW (Figure 2A). The reduction of nAb titer against the beta VoC was evident in all disease categories. RBD IgG arbitrary units also showed a correlation with previous disease severity but the difference in peak IgG binding between Wuhan-Hu-1 spike and VoCs antigens within each HCW category was not significant (Figure 2B).

**Figure 2.**
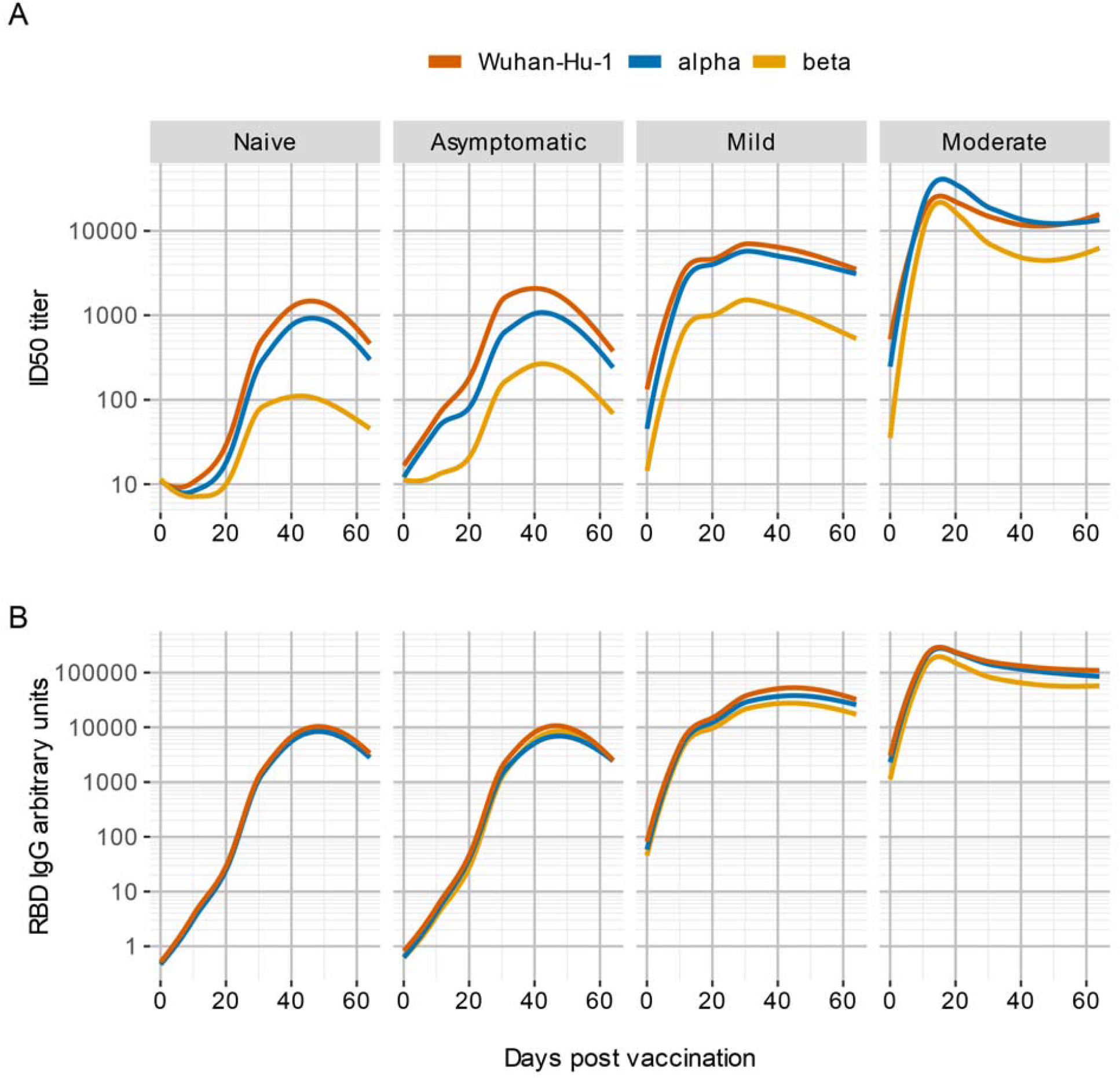
Post BNT162b2 vaccination, antibody levels to spike antigens of SARS-CoV-2 and its variants are correlated to previous disease severity. The line plots show the moving average of antibody levels against the indicated SARS-CoV-2 variants after fitting of a LOESS polynomial regression. Subjects are stratified according to symptoms after SARS-COV-2 infection. Antibody responses to Spike antigens of SARS-CoV-2 beta VoCs post BNT162b2 vaccination show significant differences in nAbs titer **(A)** but not in IgG binding to the RBD **(B)**.

### An early boost of HKU1 IgGs post-vaccination is associated with a rapidly decreasing Ab response against SARS-CoV-2 antigens in COVID-19 naïve subjects

Vaccination induced a large, early boost of IgG against seasonal betacoronaviruses in a fraction of HCW. This rapid increase of IgG binding to the Spike S2 antigens of OC43 and HKU1 seasonal coronaviruses occurred already at day 10 after the first vaccination in 7 out of 13 naïve HCW and 3 of 6 HCW with prior COVID-19 (all of whom SARS-CoV-2 Abs negative at baseline) (Figure 3, A and B). In all HCW who presented with SARS-CoV-2 Abs at baseline, the Ab response against Wuhan-Hu-1 spike antigens at day 10 invariably exceeded that against seasonal betacoronaviruses S2 (Figure 3C). The expanded IgG response to HKU1 and OC43 S2 antigens appeared to be minimally if at all cross-reactive with SARS-CoV-2 S2

**Figure 3.**
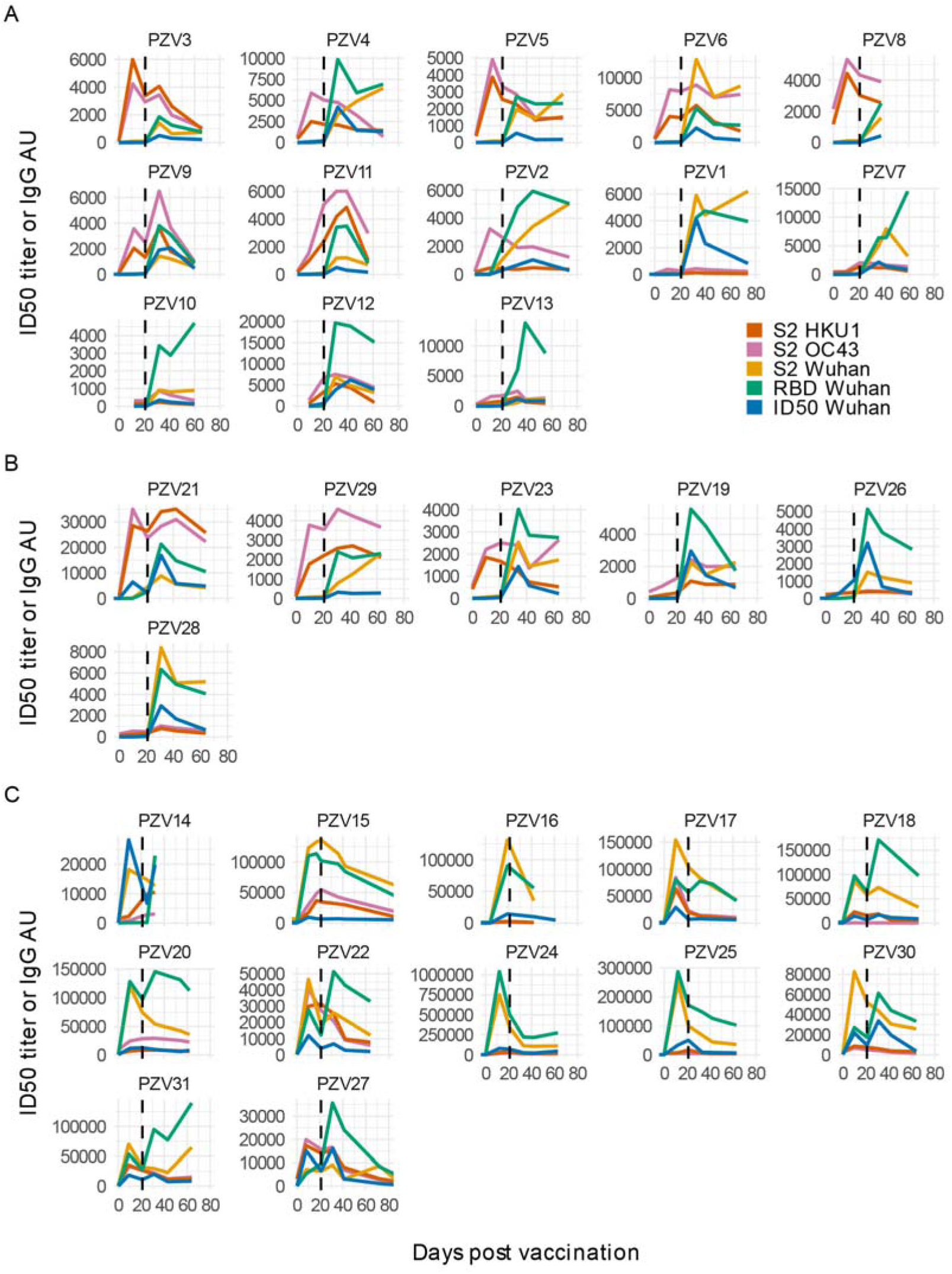
BNT162b2-Comirnaty vaccination can induce a large, early boost of antibodies against seasonal betacoronaviruses in COVID-19 naïve subjects. Line plots of ID50 nAbs and RBD IgG arbitrary units (AU) to SARS-CoV-2 Wuhan-Hu-1 and S2 spike proteins of SARS-CoV-2, OC43 and HKU1 betacoronaviruses at sequential time-points after vaccination. Vaccinees are stratified as: **(A)** subjects naïve for SARS-CoV-2 infection (n=13), **(B)** subjects with prior confirmed COVID-19 presenting at vaccination either without Wuhan-Hu-1 nAbs and RBD IgGs (n=6) and **(C)** with prior COVID-19 and SARS-CoV-2 antibodies at baseline (n=12). The vertical dashed line indicates the 2nd vaccine jab.

A high titer of HKU1 IgGs at day 10 was associated with lower peak values of SARS-CoV-2 antibodies and their quicker decline during follow-up (two-way repeated measures ANOVA p <0.015). At day 42, nAbs against the beta VoC dropped below detection limit in 4 out of 7 naïve subjects with an early boost of HKU1 antibodies compared to 1 of 6 of those without (Figure 4A). A significant post peak reduction of RBD IgG arbitrary units during follow-up was also present in HCW with an early boost of HKU1 antibodies compared with those without (two-way repeated measures ANOVA p <0.05) (Figure 4B).

**Figure 4.**
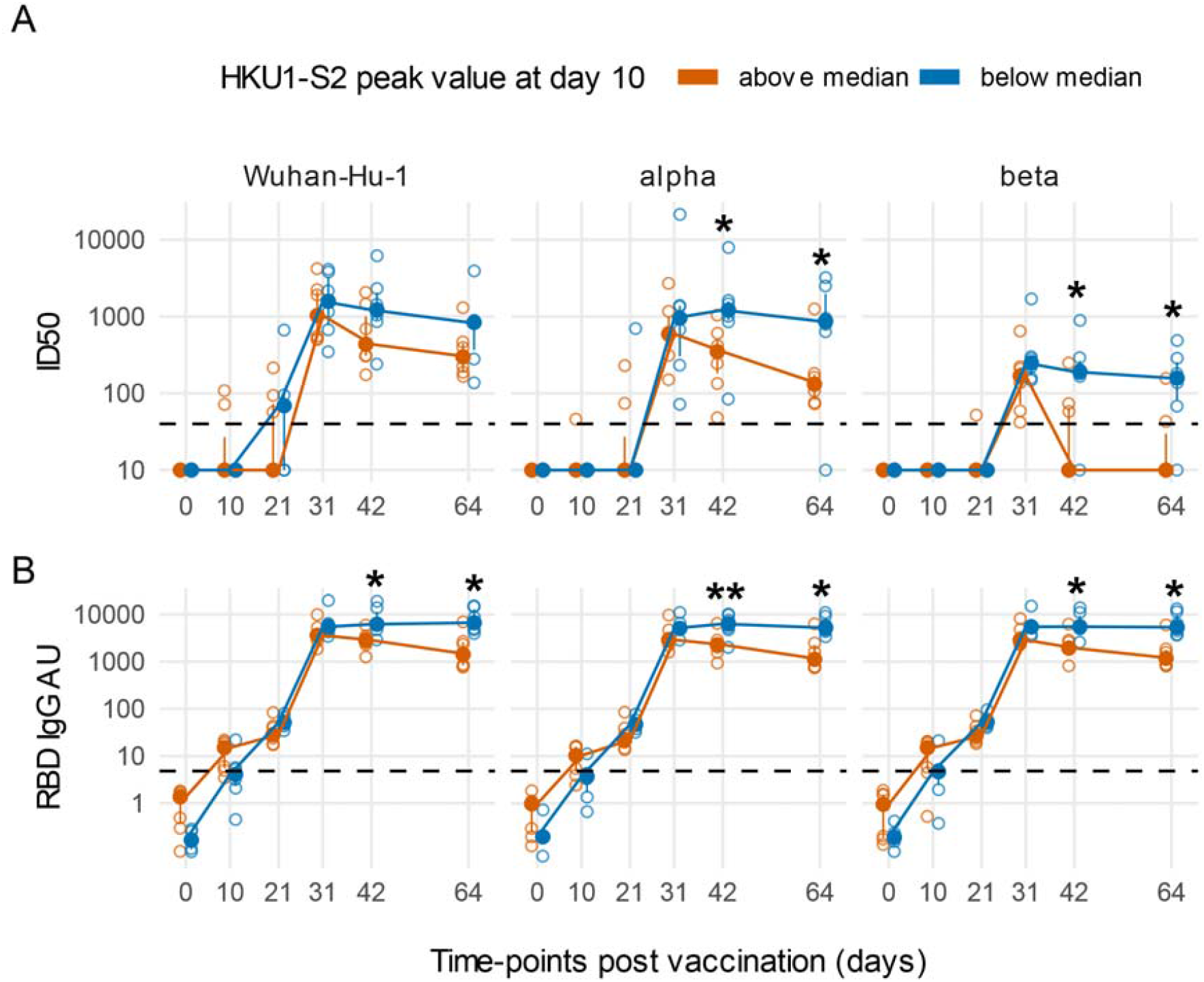
An early boost of HKU1 IgGs post-vaccination is associated with a rapidly decreasing Ab response against SARS-CoV-2 in COVID-19 naïve subjects. Line plots of Wuhan-Hu-1 and VoCs ID50 nAbs **(A)** and RBD IgG AU **(B)**. Subjects naïve for COVID-19 were stratified as above or below the median of HKU1 S2 IgGs at day 10 post-vaccination. Filled circles and bars represent the median ± inter quartile range at each time-point, empty circles show each individual measurement. The horizontal dashed lines stand for the threshold for positivity. The asterisks indicate statistically significant differences in mean ID50 nAbs or RBD IgG AU at the corresponding time-points between subjects with or without an early boost of seasonal coronaviruses responses after vaccination (two-way repeated measures ANOVA, * p < 0.05, ** p< 0.01).

To exclude a generalized impact of vaccination on pre-existing antibody responses we measured IgG binding to the HA antigen of the 2009 pandemic H1N1 flu virus in COVID-19 naïve HCW. HA antibodies showed modest fluctuations during follow-up, which were not synchronous with those against betacoronaviruses’ antigens (Supplemental Figure 3).

Overall, our data indicate that vaccinees naïve for COVID-19 frequently show a rapid drop below detection limit of antibodies that cross-neutralize VoCs. Moreover, a similar profile and kinetics of post-vaccination antibodies was seen in some vaccinees who recovered from prior asymptomatic or mild COVID-19 and were Wuhan-hu-1 nAbs negative at the time of vaccination. This was at variance with the vaccination outcome in vaccinees with prior COVID-19 with detectable Wuhan-hu-1 nAbs at baseline, who had a neutralizing response an order of magnitude greater and sustained throughout follow-up. The observed rapid decrease of antibodies able to neutralize VoCs in vaccinees either naïve for SARS-CoV-2 infection or who experienced asymptomatic/mild COVID-19 should inform the debate on strategies aimed at boosting protection from VoCs infection.

Secondly, our study shows that the nAb response kinetics is not mirrored by that of IgG binding to the RBD. IgG binding to the Wuhan-hu-1 and VoC RBDs is similar and shows only a modest drop by the end of the follow-up. This observation agrees with the recent evidence that many RBD IgGs in vaccinees are non-neutralizing and that several nAbs target the spike protein NTD rather than the RBD (Amanat et al., 2021). Therefore, caution should be exercised in using RBD antibodies as proxy for VoC neutralization or as a surrogate marker to predict vaccine effectiveness. Moreover, current large-scale screening immunoassays measuring RBD antibodies are essentially all based on the Wuhan-Hu-1 sequence. Our study suggests that simply switching the antigen RBD sequence to that of VoCs would not lead to an efficient detection of differences in neutralizing activity among screened sera.

Finally, our data shows that BNT162b2 vaccination frequently induces an early boost of antibodies to the S2 subunit of OC43 and HKU1 seasonal betacoronaviruses. This boost was rapid and large in >50% of vaccinees naïve for SARS-CoV-2 infection. Importantly, the rapid increase of IgG to HKU1 at 10 days post-mRNA vaccination was associated with a faster decrease and subsequent loss of SARS-CoV-2-specific antibodies. This was particularly evident against VoCs, with a loss of neutralization against the beta VoC in the majority of naïve vaccinees who experienced this early boost of seasonal coronaviruses antibodies.

We previously described an increase of antibodies against HKU1 and OC43 Spike antigens in a cohort of symptomatic COVID-19 patients shortly after disease onset (Secchi et al., 2020). In those patients, HKU1 and OC43 antibody levels were significantly correlated with those of SARS-CoV-2 specific antibodies during the first 3-4 weeks from symptoms onset. In addition to similar observations (Kaplonek et al., 2021), the existence of cross-reactive antibodies between SARS-CoV-2 and seasonal betacoronaviruses has been further demonstrated by the isolation of cross-reactive monoclonal antibodies from germinal centers of vaccinated individuals (Amanat et al., 2021). The coding sequence of these cross-reactive monoclonal had more extensive somatic mutations compared to non-cross-reactive ones, signaling a likely derivation from memory responses pre-existing vaccination. Moreover, these findings are consistent with the existence of betacoronaviruses cross-reactive T cells in subjects that were either naïve for SARS-CoV-2 infection or had COVID-19 (Woldemeskel et al., 2020; Braun et al., 2020; Grifoni et al., 2020).

It is relatively straightforward to speculate that the observed early rapid expansion of HKU1 and OC43 IgG antibodies post-vaccination could be attributed to the presence of both elevated pre-existing memory B cells to seasonal coronaviruses and cross-reactive helper T cells recognizing peptide epitopes in the Spike S2 subunit, due to its relatively high homology between coronaviruses. However, it is unclear how the expansion of memory responses to other betacoronaviruses Spike S2 might subsequently impact on SARS-CoV-2 nAbs development. The S2 subunit is rarely if ever directly targeted by SARS-CoV-2 nAbs (Graham et al., 2021). It is therefore unlikely that an early deviation of antibody responses towards the S2 of seasonal betacoronaviruses rather than that of SARS-CoV-2 might lead to a direct decrease of nAbs. Furthermore, our data suggest also a partially generalized reduction of SARS-CoV-2 specific antibodies including binding IgG against the spike RBD in naïve subjects with an early reactivation of seasonal betacoronaviruses B cell memory. Possibly, this observation might support the concept of a preferential and synchronous expansion of a subset of pre-existing and cross-reactive T cells recognizing S2 rather than S1 epitopes.

The persistence of detectable nAbs is key for protection from SARS-CoV-2 infection. However, modelling studies based on immunological data from mild and moderate COVID-19 patients suggest that nAbs elicited from less severe natural infection is unlikely to provide long-term protection (Wheatley et al., 2021). Our own data suggests that the re-expansion of pre-existing humoral memory to seasonal betacoronaviruses might dampen the SARS-CoV-2 neutralization response induced by vaccination. This implies that in a consistent fraction of vaccinees naïve for COVID-19 the duration of protection from subsequent re-infection would be shortened. Additionally, it might be speculated that this mechanism might be at play also in some individuals who experienced an asymptomatic infection, leading to the discrepancies in the persistence of SARS-CoV-2 nAbs that we observed.

While the effectiveness of the immunization with current mRNA vaccines is undisputable in reducing both infection and disease severity (Haas et al., 2021), public health policies must adapt to rapidly emerging VoCs. The choice of diagnostic tests, vaccine formulations and vaccine deployment will benefit from investigating further the mechanisms influencing the response to vaccination, particularly in view of the ongoing development of pan-coronavirus vaccines (Shiakolas et al., 2021).

## Materials and methods

### Study approval

All procedures performed in studies involving human participants were in accordance with the ethical standards of the institutional and/or national research committee and with the 1964 Helsinki declaration and its later amendments or comparable ethical standards. Written informed consent was obtained from all individual participants included in the study.

The study subjects belonged to the IRCCS Ospedale San Raffaele COVID-19 cohort study COVID-BioB (registered as ClinicalTrialsgov-NCT04318366) and approved by the IRCCS Ospedale San Raffaele Ethics Review Board (protocol 68/INT/2020) and the COVID-BioB related immunological sub-studies “Role of the immune response in the infection with SARS-CoV-2 and in the pathogenesis of COVID-19” (protocol number 34/int/2020).

### Study population

The study subjects included 31 hospital health care workers (HCW) of the IRCCS Ospedale San Raffaele, who received their first dose of the Pfizer-BioNtech mRNA vaccine BNT162b2 – Comirnaty between January 7th and March 5th 2021. All but two subjects received their booster vaccination after 21 days according to Italian national guidelines (https://www.gazzettaufficiale.it/atto/serie_generale/caricaDettaglioAtto/originario?atto.dataPubblicazioneGazzetta=2021-03-24&atto.codiceRedazionale=21A01802&elenco30giorni=false) (Supplementary Table 1). Thirteen were naïve to COVID-19 at vaccination, while 18 had a previous SARS-CoV-2 infection either during the first pandemic wave between February to March 2020 (n=13, median interval from symptoms onset to first jab 302.5 days or during the second wave between October to November 2020 (n=5, median interval from symptoms onset to first jab 76 days). Four individuals with COVID-19 with a confirmed negative nasopharyngeal swab post-infection, tested positive at a routine in-hospital screen 1.5 to 8 months later likely due to asymptomatic re-infection events.

A confirmed infection case was defined as a SARS-CoV-2 positive real-time reverse-transcriptase polymerase chain reaction (RT-PCR) from a nasopharyngeal swab and/or COVID-19 symptoms and/or a serologic evidence of antibody to SARS-CoV-2. The WHO classification was used to define disease severity (https://www.who.int/publications-detail-redirect/WHO-2019-nCoV-clinical-2021-1). Asymptomatic infections were captured thanks to the hospital SARS-CoV-2 testing policy, which introduced in May 2020 a serologic antibody test (LIAISON® SARS-CoV-2 S1/S2 IgG serological test, DiaSorin S.p.A., Vercelli, Italy) and in August 2020 a regular monthly or bimonthly SARS-CoV-2 nasal-pharyngeal swab test. The clinical and laboratory data were collected from medical chart review or directly by interview, crosschecked for accuracy by data managers and clinicians and entered in a dedicated electronic case record form (eCRF) developed on site for the COVID-BioB study in MySQL (V5.7.14) on Apache Tomcat (V2.4.23) platform running on windows sever 2012 R2.

Biological material of the vaccinees included a dedicated serum sample collected at first vaccination, and thereafter at 10, 21, 31, 42, 64 days (with an approximation of +- 5 days). All serum samples were coded and anonymously processed for the immunological studies.

### Construction of plasmids for lentiviral pseudotypes expression of variant spikes

A schematic representation of recombinant antigens used in this study is shown in Supplementary Fig. 1.

Plasmid pSpike-C3 expressing the codon optimized SARS-CoV-2 Spike protein open reading frame (ORF) (GenBank: NC_045512.2) containing a 21 amino acid deletion at the cytoplasmic tail (delta21) of Spike protein were previously described (Dispinseri et al., 2021). For construction of pSpike-UKC3 and pSpike-SAC3 plasmids, expressing the variant Spike ORFs with a 21 amino acid deletion at the cytoplasmic tail, a NheI/AvaI fragment of DNA was removed from Alpha (B.1.1.7 pSpike-UK) and Beta (B.1.351 pSpike-SA) plasmids and inserted into the corresponding restriction sites of pSpike-C3 plasmid, to obtain pSpike-UKC3 and pSpike-SAC3 plasmids. The B.1.1.7 pSpike-UKC3 used in these studies contained the following mutations: del69-70HV, del145Y, N501Y, A570D, D614G, P681H, T716I, S982A, D1118H. The B.1.351 pSpike-SAC3 used in these studies contained the following mutations: L18F, D80A, D215G, del242-244LAL, R246I, K417N, E484K, N501Y, D614G, A701V (Supplementary Figure 1).

### Production of lentiviral vector expressing luciferase (LV-Luc) and pseudotyped with Spike variants

293T Lenti-X human embryonic kidney cell line (Clontech, Mountain View, CA, USA) was used for production of LV-Luc pseudotyped with Spike variants by transient transfection as previously described (Dispinseri et al., 2021). Briefly, 293T Lenti-X cells (3.5×106 cells) were seeded on 10 cm Petri dishes (Corning Incorporated - Life Sciences, Oneonta, NY, USA) and transiently transfected with plasmids pGAE-LucW, pADSIV3+ and pseudotyping plasmid (pSpike-C3, pSpike-UKC3, pSpike-SAC3) using the JetPrime transfection kit (Polyplus Transfection Illkirch, France) following the manufacture’s recommendations using a 1:2:1 ratio (transfer vector: packaging plasmid: Spike plasmid). At 48 h post transfections, culture supernatants containing the LV-Luc pseudotypes (LV-Luc/Spike-C3, LV-Luc/Spike-UKC3 and LV-Luc/Spike-SAC3) were collected and stored in 1 mL aliquots at -80°C until use.

### Lentiviral vector-based SARS-CoV-2 neutralization assay

LV-Luc preparations were titered on VeroE6 cells (African green monkeys, epithelial kidney) and dilutions providing 150,000-200,000 relative luciferase units (RLU) were used in the neutralization assay, as previously described (Dispinseri et al., 2021). Briefly, heat-inactivated serum serial 3-fold dilutions starting from the 1/40 dilution were incubated in duplicate with the LV-Luc for 30 min at 37°C in 96-well plates, and thereafter added to VeroE6 cells at a density of 20.000 cells/well. After 48 h luciferase expression was determined with a luciferase assay system (Bright-Glo, Promega) and measured in a Mitras luminometer (Berthold, Germany). The 50% inhibitory serum dilution (ID50) was calculated with a linear interpolation method using the mean of the duplicate responses (Fenyö et al., 2009). Neutralization was expressed as the reciprocal of the serum dilution giving 50% inhibition of RLU compared to the mean of the virus control wells. An ID50 below 1/40 serum dilution was considered negative and a value of 10 ascribed for statistical analysis.

### IgG binding antibody Luciferase Immuno Precipitation System (LIPS) assay

Using LIPS (Secchi et al., 2020; Burbelo et al., 2005) we measured IgG binding to recombinant nanoluciferase tagged antigens corresponding to SARS-CoV-2 alpha, beta and gamma spike RBD and S2 domains, to HKU1 and OC43 seasonal betacoronaviruses spike S2 domains, and hemagglutinin (HA) antigen of the pandemic Ca2009 H1N1 influenza virus as previously described (Secchi et al., 2020; Lind et al., 2021) (Supplemental Figure 1). Briefly, we cloned recombinant nanoluciferase-tagged monomeric or multimeric antigens and expressed them by transient transfection into Expi293F™ cells (Expi293™ Expression System, Thermo Fisher Scientific Life Technologies, Carlsbad, CA, USA). For LIPS, we incubated in liquid phase each antigen with test serum (1ul) for 2 h and then captured immune-complexes with rProtein A-sepharose. After washing (5 times) the sepharose pellets, we quantified bound IgG by measuring the recovered luciferase activity in a Berthold Centro XS3 luminometer (Berthold Technologies GmbH & Co. KG, Bad Wildbad, Germany) using the MikroWin version 5.22 software. We then converted raw data into arbitrary units (AU), for SARS-COV-2 specific antibodies we used a local positive index serum that exhibited closely similar binding to each of the VoCs, for HKU1 and OC43 specific antibodies we used two local index sera with strong antibody binding to the respective betacoronavirus antigen.

### Statistical analysis

We performed statistical analyses using the R software version 3.4.1 (R Core Team, 2020). We reported the median with either 95%CI, range, or inter-quartiles range (IQR) for continuous variables and frequency or percent % frequency for categorical variables. We used repeated measure ANOVA to compare continuous variables over time. Two-tailed p values are reported, with p value <0.05 indicating statistical significance.

## Data Availability

Raw data that support the findings of this study are available upon reasonable request.

## Data Availability

Raw data that support the findings of this study are available upon reasonable request.

## Acknowledgements

The work performed at IRCCS Ospedale San Raffaele (OSR) was funded by Program Project COVID-19 OSR-UniSR and Ministero della Salute (COVID-2020-12371617). The work by Viral Evolution and Transmission Unit, OSR, Istituto Superiore di Sanità (ISS) and Imperial College was funded by EAVI2020, the European Union’s Horizon 2020 research and innovation programme under grant agreement No. 681137. ISS received support in part by NATO multi-year Project No. G5817 “New and Validated Tools for the Diagnosis and follow-up of SARS-CoV-2 Infected Individuals” and by ISS internal funds. We thank Fondation Dormeur, Vaduz for the donation of laboratory instruments relevant to this project to the Viral Evolution and Transmission Unit at OSR and to the National Center for Global Health at the Istituto Superiore di Sanità. A special acknowledgement goes to the COVID-BioB team and health-care workers at OSR that made this work possible. The sponsors had no role in study design; in the collection, analysis, and interpretation of data; in the writing of the report; and in the decision to submit the paper for publication.

## Author notes

GS, VL, ACara, PC conceived and designed the study. GS, SD, VL wrote the first draft with contributions from ACara, DN, LP. VL, IM, GS did the statistical analysis. SD, MT, CB, EB, MP, PMK, ACan, MB, and AG did the laboratory tests. SD, IM, VL, and GS did the data organization, data storage and quality control. SD, RC, RV, and CB oversaw sample handling and storage. GS, FC, and PC oversaw institutional review board submissions, approval, and clinical study oversight. GS, LP, VL, FC, ACara, DN provided funding. VL, GS, DN, ACara, IM, SD prepared the manuscript for submission. VL and GS jointly supervised this work. All authors reviewed the manuscript for intellectual content and assisted in data interpretation. All authors confirm that they had full access to all the data in the study and accept responsibility to submit for publication.

