## Supplemental figures 1-3 and tables 1-2 for "Seasonal betacoronavirus antibodies expansion post BNT161b2 vaccination associates with reduced SARS-CoV-2 VoCs neutralization"

Supplemental figure 1

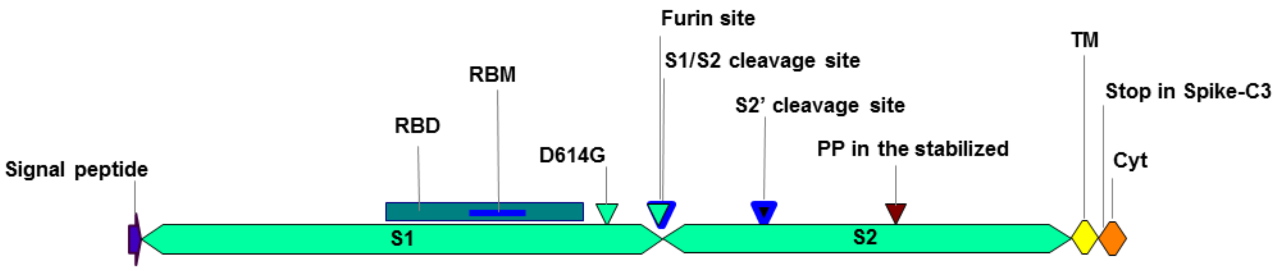


#### Nab assay antigens

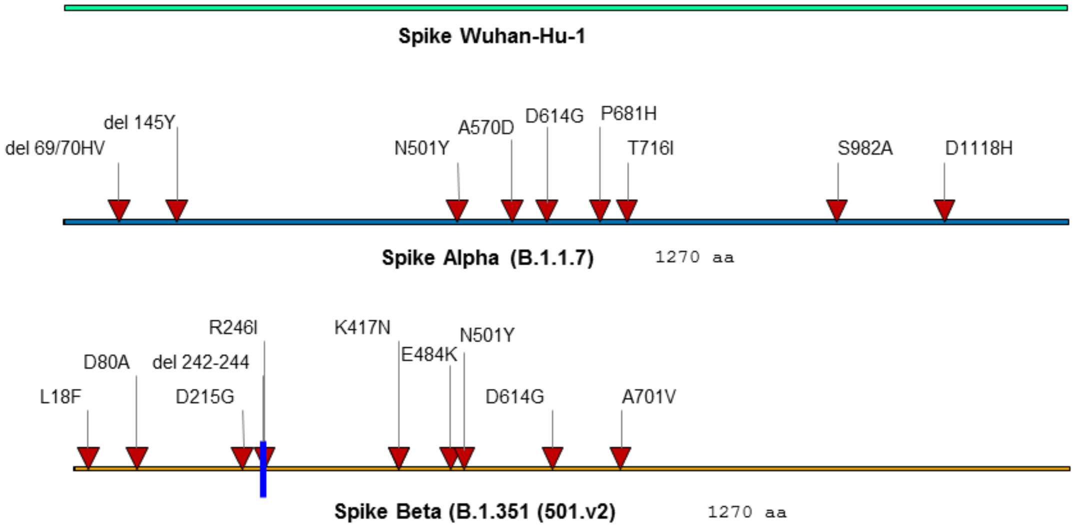


#### LIPS antigens

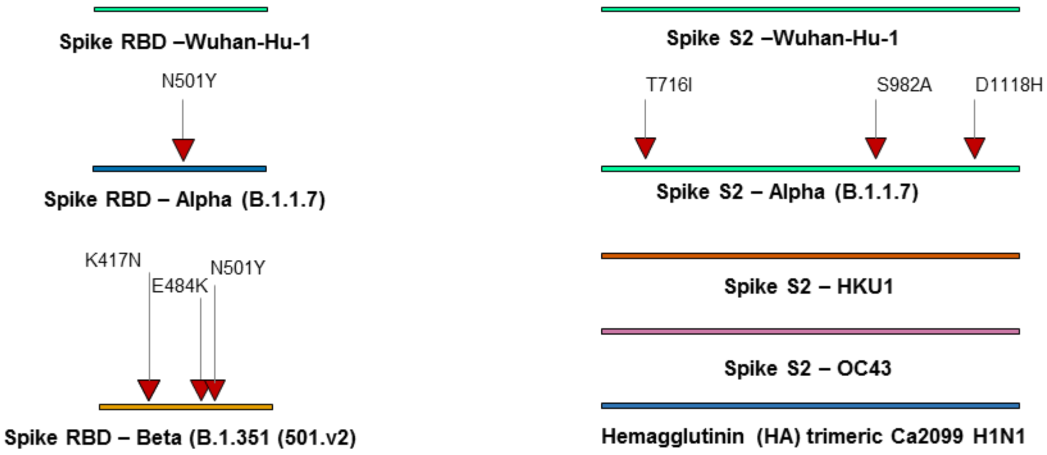


**Schematic representation of SARS-CoV-2 constructs used in Nabs and LIPS assays**

Supplemental figure 2

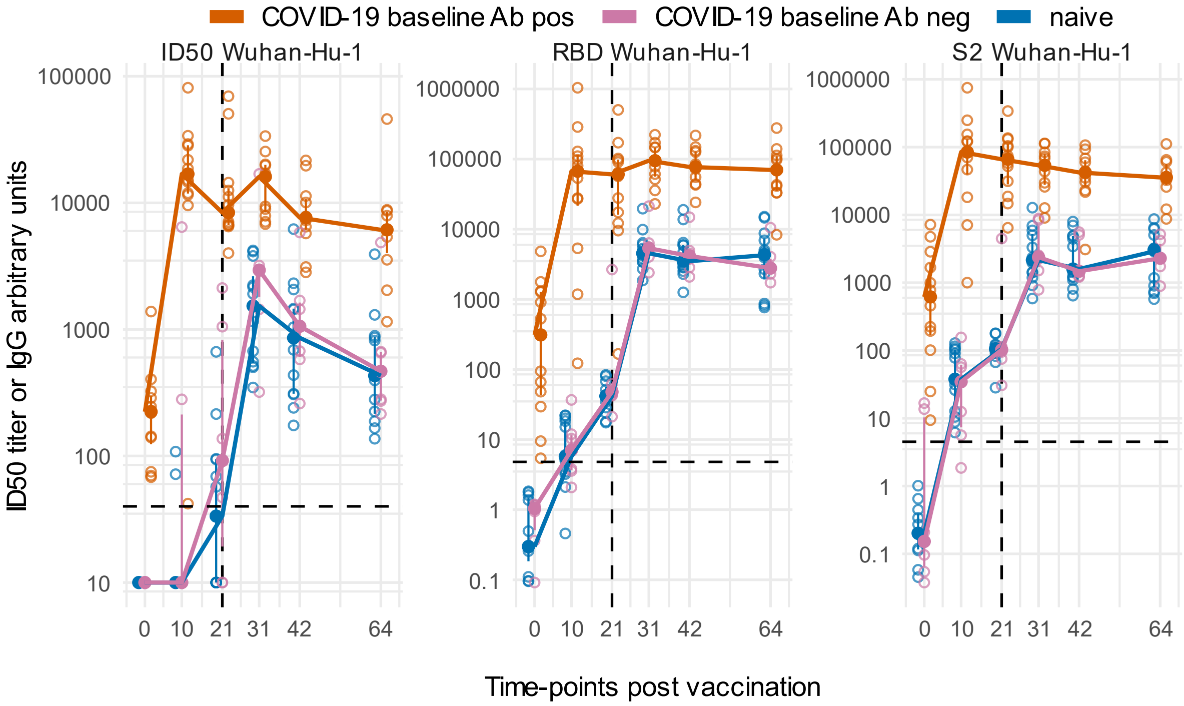


**Antibody responses against Wuhan in BNT162b2 vaccinees stratified by SRAS-CoV-2 Ab status at baseline** Line plots show the temporal profile post BNT162b2 vaccination of ID50 titers or IgG arbitrary units against the Spike RBD or S2 domains. Vaccinees are stratified according to previous infection with SARS-CoV-2 into naïve or with previous COVID-19 either without or with SARS-CoV-2 neutralizing and RBD antibodies at baseline before vaccination (COVID-19 baseline Ab neg or COVID-19 baseline Ab pos, respectively). Filled circles with error bars correspond to median ± IQR at the indicated timepoints. Empty circles correspond to individual subject values. Horizontal dashed lines indicate the respective assay threshold for positivity. The vertical dashed line indicates the second vaccine jab timepoint.

Supplemental figure 3

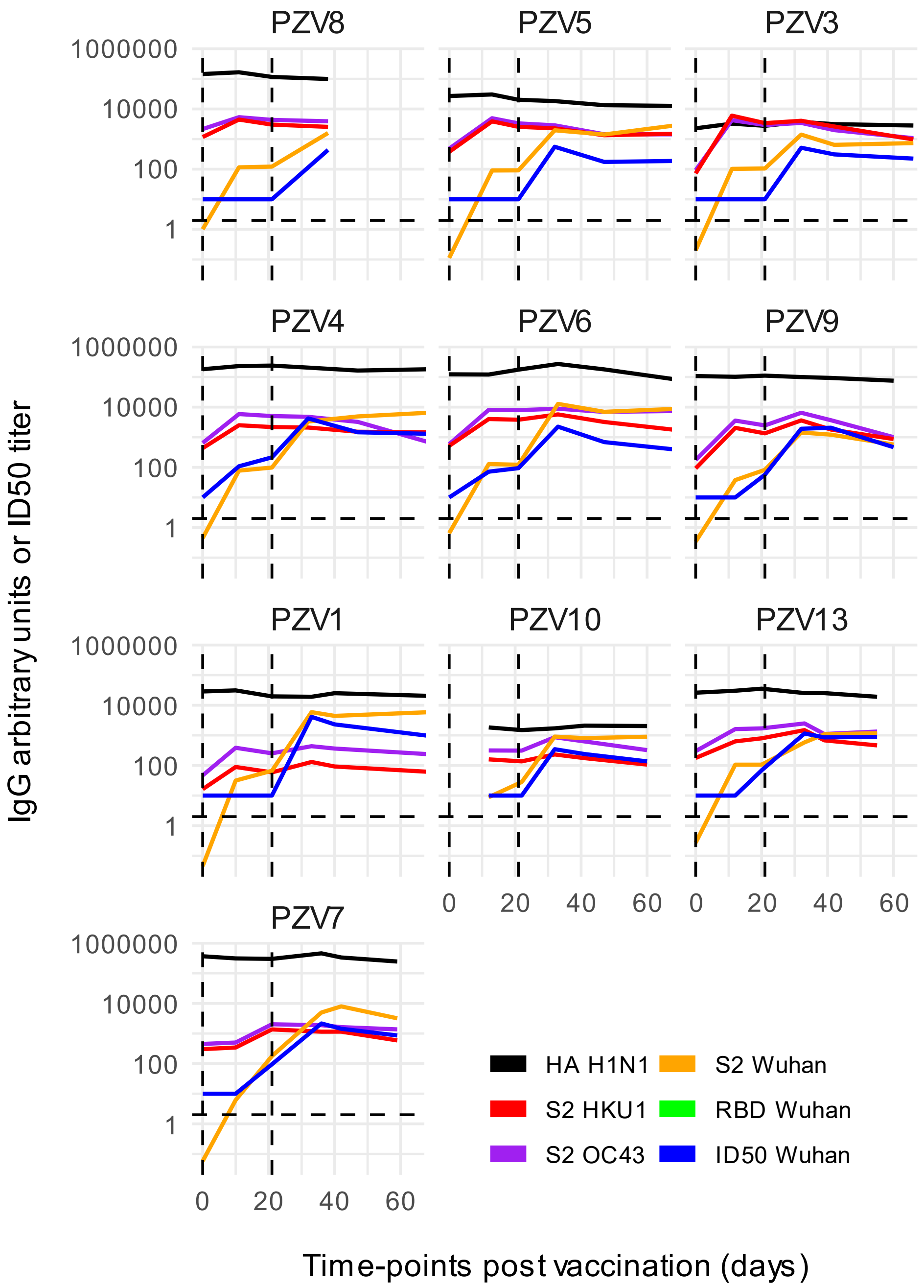


**The antibody response to the pandemic H1N1 flu virus HA antigen is not affected by vaccination.** The line plots show the temporal profile post BNT162b2 vaccination of antibody levels in binding or neutralization assays in a selection of SARS-CoV-2 naïve BNT162b2 vaccinees. HA antibodies show modest fluctuation over time that are not synchronous with hose against betacoronaviruses' antigens. The dashed line indicates the second BNT162b2 jab timepoints.

### Supplemental Table 1. Demographics and clinical characteristics of study subjects

| Subject ID | Sex | Age at vaccination (years) | N. of vaccine shots | COVID-19 | Symptoms category at first COVID-19 | Symptoms category at second COVID-19 | Days from symptoms onset to first vaccination (range) | SARS-CoV-2 alpha nAbs and RBD IgG at baseline |
|---|---|---|---|---|---|---|---|---|
| PZV1 | F | 56-60 | 2 | Naïve | | | | No |
| PZV2 | M | 61-65 | 2 | Naïve | | | | No |
| PZV3 | F | 26-30 | 2 | Naïve | | | | No |
| PZV4 | F | 26-30 | 2 | Naïve | | | | No |
| PZV5 | M | 31-35 | 2 | Naïve | | | | No |
| PZV6 | M | 31-35 | 2 | Naïve | | | | No |
| PZV7 | F | 56-60 | 2 | Naïve | | | | No |
| PZV8 | F | 56-60 | 2 | Naïve | | | | No |
| PZV9 | F | 36-40 | 2 | Naïve | | | | No |
| PZV10 | F | 41-45 | 2 | Naïve | | | | No |
| PZV11 | M | 31-35 | 2 | Naïve | | | | No |
| PZV12 | F | 46-50 | 2 | Naïve | | | | No |
| PZV13 | M | 36-40 | 2 | Naïve | | | | No |
| PZV14 | F | 26-30 | 2 | First wave | asymptomatic | | unknown | Yes |
| PZV15 | F | 56-60 | 1 | First wave | moderate | | 350 -400 | Yes |
| PZV16 | F | 46-50 | 1 | First wave | mild | | 250-300 | Yes |
| PZV17 | F | 41-45 | 2 | First wave | mild | | 300-350 | Yes |
| PZV18 | F | 51-55 | 2 | First wave | mild | | 250-300 | Yes |
| PZV19 | F | 51-55 | 2 | First wave | asymptomatic | | 300-350 | No |
| PZV20 | M | 31-35 | 2 | First wave | mild | | 300-350 | Yes |
| PZV21 | F | 66-70 | 2 | First wave | mild | | 300-350 | No |
| PZV22 | F | 56-60 | 2 | First wave | mild | | 300-350 | Yes |
| PZV23 | F | 46-50 | 2 | First wave | asymptomatic | | >250 | No |
| PZV24 | F | 51-55 | 2 | First wave – Second wave | moderate | asymptomatic | 300-350 | Yes |
| PZV25 | F | 46-50 | 2 | First wave – Second wave | mild | asymptomatic | 250-300 | Yes |
| PZV26 | M | 31-35 | 2 | First wave – Second wave | asymptomatic | asymptomatic | >200 | No |
| PZV27 | F | 46-50 | 2 | Second wave - 2020 | mild | mild | 50-100 | Yes |
| PZV28 | F | 51-55 | 2 | Second wave | asymptomatic | | 50-100 | No |
| PZV29 | F | 26-30 | 2 | Second wave | mild | | 50-100 | No |
| PZV30 | M | 61-65 | 2 | Second wave | mild | | 50-100 | Yes |
| PZV31 | F | 31-35 | 2 | Second wave | mild | | 50-100 | Yes |

#### Supplemental Table 2. Peak values of SARS-CoV-2 antibodies post vaccination

| | Geometric mean at day 31 post vax | | Geometric mean ratio |
|---|---|---|---|
| | HCW with prior COVID-19 and detectable SARS-CoV-2 abs at baseline before vaccination | HCW without detectable SARS-CoV-2 abs at baseline before vaccination | HCW with baseline Abs / HCW without |
| **Wuhan-Hu-1 ID50 titer** | 13613 | 1607 | 8.5 |
| **Wuhan-Hu-1 RBD IgG arbitrary units** | 84173 | 5162 | 16.3 |
| **Wuhan-Hu-1 S2 IgG arbitrary units** | 43733 | 2578 | 17.0 |
